# Chlamydia and Gonorrhea Incidence and Residential Segregation: US Spatiotemporal Patterns (2013–2021)

**DOI:** 10.1101/2025.06.02.25328114

**Authors:** M. Naser Lessani, Arielle N’Diaye, Zhenglong Li, Xiaoming Li, Shan Qiao

## Abstract

**Objective:** Investigate how residential segregation is associated with chlamydia and gonorrhea incidence between 2013 and 2021.

**Methods:** National-level secondary US data from 2013-2021 from the Centers for Diseases Control and Prevention Sexually Transmitted Infection surveillance dataset, American Community Survey, and Racial Segregation Index were analyzed using the Generalized Estimating Equation, and spatial regression. Analysis was divided into two periods (2013–2019; 2020–2021) to account for COVID-19 disruptions. Residential segregation was measured by dissimilarity index categorized into reference (<□0.25), moderate (0.26–0.50), high (0.51–0.75), and extreme (>□0.75) levels. Primary outcome measures were chlamydia and gonorrhea incidence rates. Residential segregation was the key independent variable with other social determinants of health covariates. 3058 counties within the contiguous United States were included within this study. Counties with missing data, and not within the contiguous United States were excluded.

**Results:** For chlamydia, from 2013–2019, segregation coefficients (i.e.,13.77 and 15.84 for moderate and high segregation) indicate that greater residential segregation is associated with higher chlamydia incidence rates (P<.0001). From 2020–2021, these coefficients increased (from 13 and 15 to 28.25 and 34.16), suggesting growing segregation-driven disparities. Gonorrhea followed a similar trend, with the coefficients increasing from 0.47 and 0.55 (P < .001) to 1.53 and 1.62 (P < .05), respectively. Spatial variation in the association between segregation and chlamydia incidence remained consistent, with stronger associations in the Southeastern, Midwest, and Western regions. Spatial variation in the association between segregation and gonorrhea incidence was more pronounced in the South and parts of the Midwest, with weaker associations in some Northern and Western regions.

**Conclusions:** Residential segregation remained a substantial driver of chlamydia and gonorrhea transmission. The spatial patterns varied over time for both diseases. Further research should extend post-COVID-19 analysis to assess evolving relationships between residential segregation and STI incidence across U.S. regions.

**Key points:** **What is already known on this topic:** Chlamydia and gonorrhea in 2023 were the most reported sexually transmitted infections in the United States, disproportionately affecting Black Americans.

**What this study adds:** This study found that racial residential segregation was a substantial driver of chlamydia and gonorrhea transmission, especially during the COVID-19 pandemic.

**How this study might affect research, practice or policy:** Study findings suggest that interventions aiming to reduce chlamydia and gonorrhea incidence rates in the United States should also include intervention activities that address adversities associated with racial residential segregation.

## 1. INTRODUCTION

Chlamydia and gonorrhea are two sexually transmitted infections (STI) that when left untreated, can result in complications like secondary infections and infertility (1). In 2023, chlamydia and gonorrhea were the top two most common STIs in the US (2). Among reported cases of chlamydia and gonorrhea, health disparities exist across gender, age, race/ethnicity and geographic regions (2, 3). Social segregation, defined as the separation of different populations within schools, workplaces, social networks, and living environments is a key driver of chlamydia and gonorrhea transmission disparities among racial minority populations (4–6). Residential segregation (i.e., the race/ethnicity-based separation of where people live) contributes to adverse health outcomes by concentrating deleterious determinants of health within living environments (e.g., inadequate housing, low quality schools, unemployment, pollution, inaccessible health care) (4, 7, 8). Black Americans are disproportionately affected by residential segregation and face heightened risks for STI acquisition due to associated environmental factors like sex-ratio imbalances, higher incarceration rates, and smaller sexual networks (6–9).

Scholars note that there is a lack of studies examining multiple indicators of socioeconomic disadvantage such as educational attainment, occupation, and health insurance status (6, 10). Though studies have used geospatial analyses to study the association of the social determinants of health (e.g., crime, race/ethnicity, and income) and chlamydia/gonorrhea transmission rates, they have been largely conducted in urban settings or concentrated settings (3, 11, 12). As such, more studies are required to understand national transmission trends across both urban and rural contexts (12). Finally, COVID-19 specific health care service interruptions and policies (e.g. shelter orders) have contributed to changes in individual-level sexual health behaviors and health seeking patterns as well as population-level STI trends (2, 13, 14). Therefore, studies are needed to explore how the COVID-19 pandemic may have affected the association between residential segregation and new STI infection rates.

To address the above-mentioned knowledge gaps, this study aims to 1) explore the associations between residential segregation and STI (i.e., chlamydia/gonorrhea) infection rates at county level in the contiguous US using national level data from 2013- 2021, and 2) to examine how these associations may be affected by the COVID-19 pandemic.

## 2. MATERIALS AND METHODS

### 2.1. Dataset and measures

We used the Centers for Disease Control and Prevention’s (CDC) surveillance data for collecting chlamydia and gonorrhea incidence rates and the ACS for salient demographic and socioeconomic information. Urbanicity characteristics were sourced from Rural-Urban Continuum Codes, which are developed by the U.S Department of Agriculture (USDA) Economic Research Service. Racial segregation index (dissimilarity index) was calculated by a research member following the established methodology (15). All study data were collected at the county level from contiguous US states from 2013 to 2021. Our analysis includes 3,058 counties excluding all counties with missing data.

Chlamydia and gonorrhea incidence rates were defined as new diagnoses numbers per 100,000 per year. Residential segregation was quantified using the dissimilarity index, which measures the evenness of distribution between White and Black populations (16). The index ranged from zero to one with lower index values indicate minimal segregation. It was categorized into four groups: reference (< 0.25), moderate, (0.26–0.50), high (0.51–0.75), and extreme (> 0.75), with values below 0.25 serving as the reference category in subsequent regression analyses.

Urbanicity was classified into three categories (i.e., urban, peri-urban, and rural) with the urban category serving as the reference group in the regression model. Additional covariates were population density, being male, being Black, educational attainment (dichotomized as having less than a high school degree versus having a bachelor’s degree or higher), poverty (percentage of population that live blow the federal poverty line), unemployment (percentage of the civilian county population that is unemployed), and health insurance coverage. We stratified age into three groups: 20–34, 35–54, and 55–64 years to address and reduce existing collinearity, as the original age group variables exhibited high multicollinearity.

Both dissimilarity index and urbanicity variables were treated as categorical variables in the analysis. The remaining variables were expressed as percentages, except the population density. After data preprocessing, the dataset’s multicollinearity was assessed using Variance Inflation Factor (VIF) measurements. Variables exhibiting high collinearity (>10) were removed.

### 2.2. Methods

Two regression models were employed: Temporal regression model, i.e., the GEE model (17), and spatial regression model, i.e., the similarity and geographically weighted regression (SGWR) model (18, 19). The GEE model provides a single coefficient value for each variable across the entire study area over time. The SGWR model offers localized coefficient estimates for each year, thereby providing deeper insight into where and how segregation influences disease rates.

#### 2.2.1. Temporal regression model

Data was first grouped into two different periods: 2013 to 2019 and 2020 to 2021 to account for the impacts of the COVID-19 pandemic. Then we used the GEE to account for the correlations within county-level data collected over time. This allowed us to have reliable population-averaged estimates. The mathematical formulation of the GEE model is:

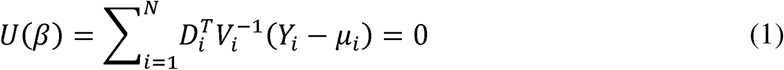

where *Y_i_* represents the response vector for the *i* th cluster, *µ_i_* is the vector of expected outcomes, 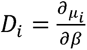 is the derivative of the mean with respect to the regression coefficients *β*, and *V_i_* shows the covariance matrix of *Y_i_* .

#### 2.2.2. Spatial regression model

Spatial regression models adhere to the assumption that everything is interrelated, and things that closer together are more closely related than those farther apart.(20) The SGWR model assessed spatial dependency by incorporating both physical proximity and variable space, which allowed us to compute a distinct coefficient for each independent variable at every location, thereby revealing how each variable varies spatially. The SGWR model can be expressed as follows:

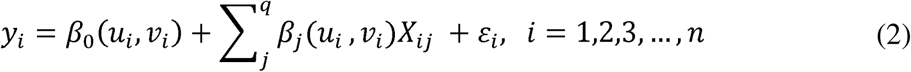

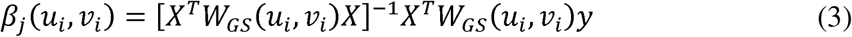

where (*u_i_,v_i_*) represents the geographical coordinates of the *i* th observation, while *e_i_* denotes the random error at that regression point. The term β_j_ (*u_i_,v_i_*) corresponds to the *j*th regression coefficient for observation *i*, highlighting that the coefficient may vary with spatial location. *w_GS_*(*u_i_,v_i_*) is the weight matrix associated with observation *i*.

## 3. RESULTS

### 3.1. Spatiotemporal distribution of chlamydia and gonorrhea

Figure 1 presents the annual aggregated incidence rates and their changes over time for contiguous states. Both chlamydia and gonorrhea in the US exhibited an overall upward trend. Chlamydia showed a steady increase from 2014 onward, with a notable decline in 2020 followed by a rebound in 2021 (Figure 1(a)), while gonorrhea incidence rates spiked significantly from 2020 to 2021 (Figure 1(b)). The Midwest and West displayed the highest chlamydia incidence rates in 2020, whereas gonorrhea incidence rates fluctuated more sharply across years (Figures 1(c) and 1(d)), with significant increases in the Midwest and Southwest. Additional state-level trends can be found in the supplementary materials (Figure S1 and S2).

**Figure 1.**
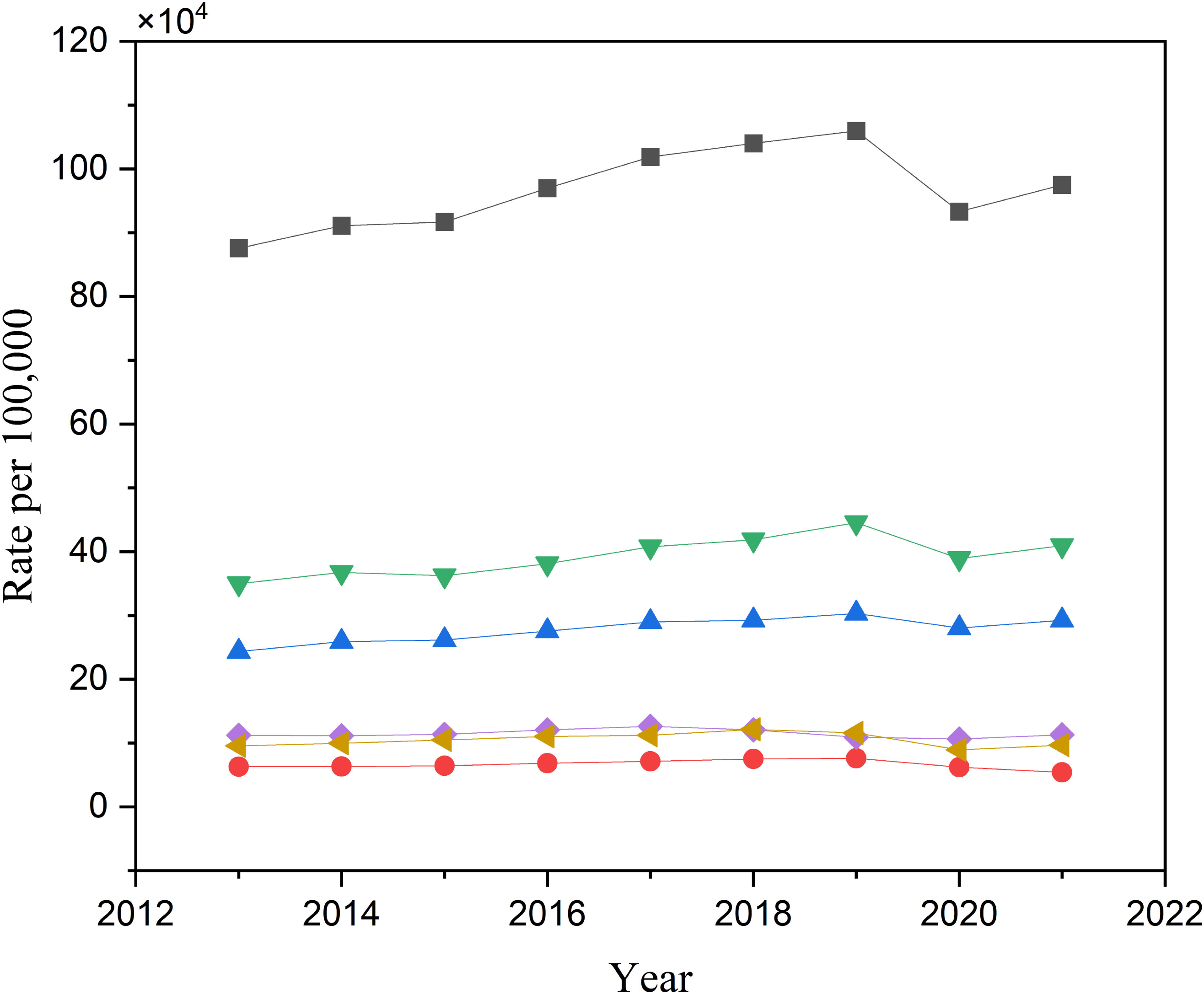
The total chlamydia incidence rate across contiguous states and regions, (b) the total gonorrhea incidence rate across contiguous states and regions. (c) and (d) the rate of change over time in incidence rate by region for chlamydia and gonorrhea, respectively. The U.S. regions are shown in (e).

Figure 2 illustrates the spatial distribution of chlamydia incidence rates from 2013 to 2021. The highest incidence rates consistently occurred in the southern U.S., particularly in the Southeastern states (Mississippi, Alabama, Georgia, South Carolina, and North Carolina), as well as parts of Arkansas, Louisiana, and Texas. Additional high-incidence clusters were observed in the Northwest (Montana, North Dakota, and South Dakota) and the Southwest (southern California, most of Arizona, and New Mexico). A notable decline was observed between 2018 and 2020 in Texas and Oklahoma, but high rates persisted elsewhere. These spatial patterns indicate that certain regions have persistently high chlamydia incidence rates, forming clear geographical clusters.

**Figure 2.**
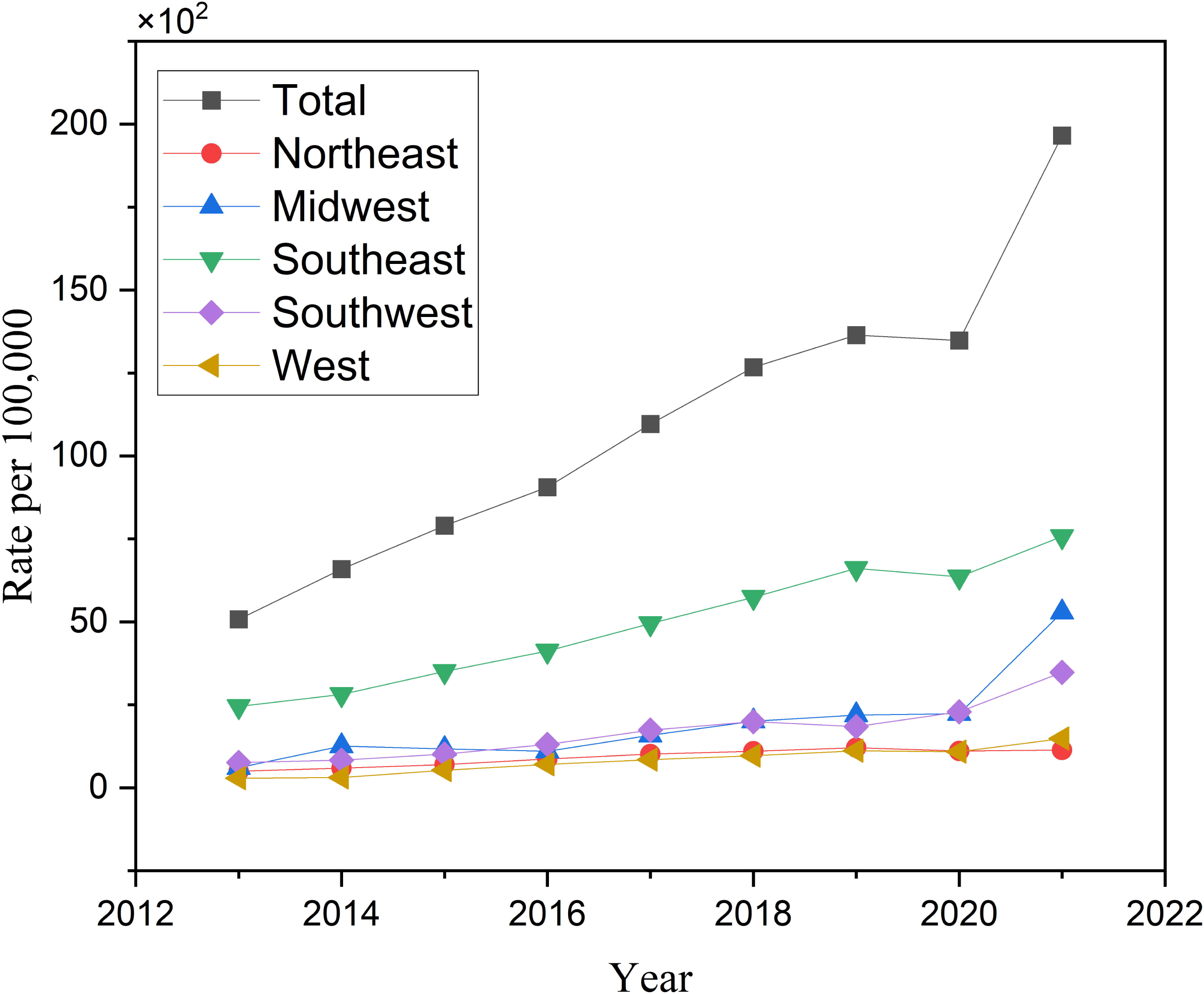
Spatial distribution of chlamydia from 2013 to 2021. The legend is displayed only on one map since the value classifications were made in the same range across all years to ensure a more comparable analysis.

Figure 3 presents the spatial distribution of gonorrhea incidence rates from 2013 to 2021. The highest rates were observed in the Southeastern U.S., particularly in Mississippi, as well as parts of Arkansas, Texas, and South Carolina. In the West and Southwest, high gonorrhea incidence was concentrated in southern California, Arizona, and New Mexico. By 2021, additional clusters had emerged in South Dakota. The spatial analysis highlights persistent geographic hotspots for gonorrhea, with concentrated regions of high incidence expanding over time, particularly after 2018.

**Figure 3.**
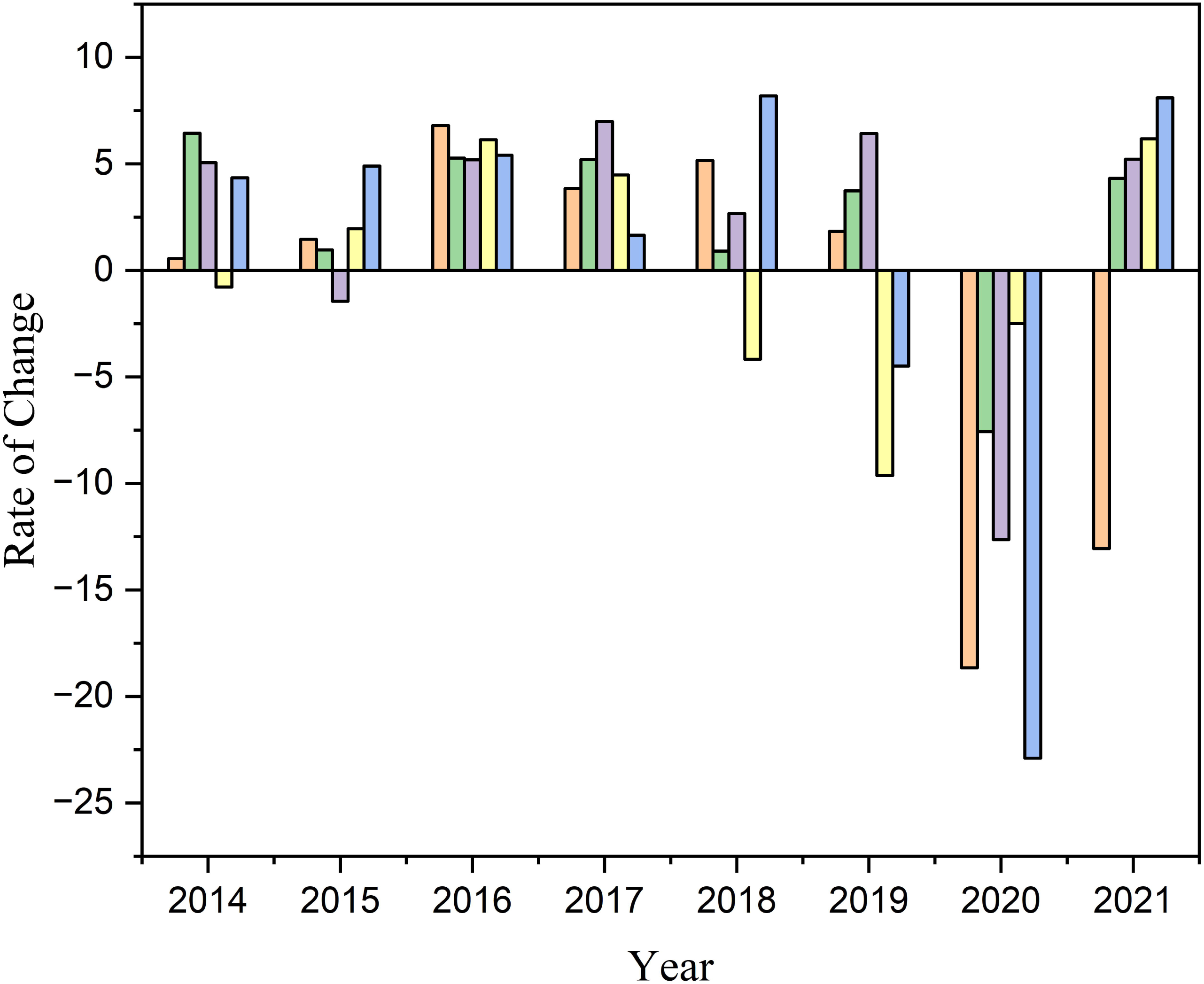
Spatial distribution of gonorrhea between 2013 and 2021. The legend is displayed only on one map since the value classifications were made in the same range across all years to ensure a more comparable analysis.

### 3.2. Temporal regression analysis

#### 3.2.1. Chlamydia

As presented in Table 1, analysis based on 2013-2019 data suggests a significant and positive association between dissimilarity index and chlamydia incidence rates controlling for all social determinants of health covariates. The coefficient of moderate and high categories for dissimilarity index was 13.77 and 15.84, respectively. This indicates that counties within this range of segregation experienced an increase of 13.77 to 15.84 chlamydia cases per 100,000 individuals compared to the reference group (index < 0.25). As segregation became more extreme, the coefficient became larger, suggesting that residential segregation significantly raised the likelihood of chlamydia infection. The results for the second period (2020-2021) were similar with a larger coefficient, which suggests that the influence of residential segregation was elevated during the COVID-19 pandemic.

**Table 1.**
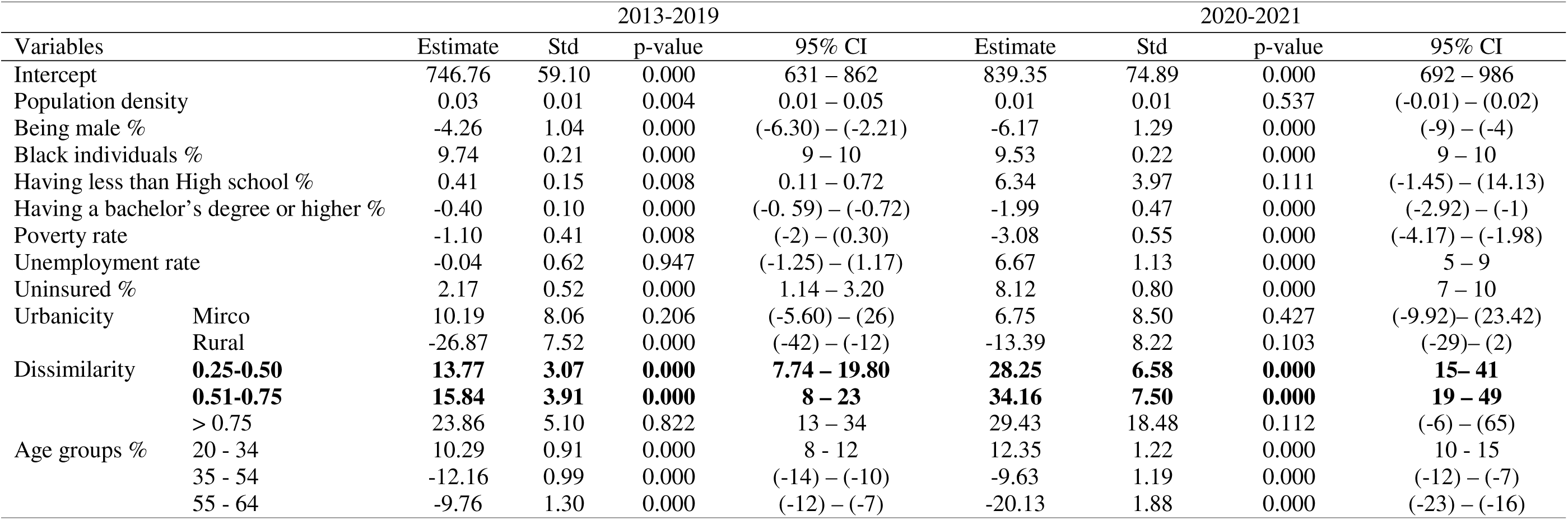
Generalized Estimating Equation’s result for Chlamydia from 2013 to 2021.

#### 3.2.2. Gonorrhea

As shown in Table 2, in the first period (2013–2019), both moderate segregation (0.47) and high segregation (0.55) showed a positive association with gonorrhea incidence rates. As the segregation index moved closer to extreme segregation (values nearer to one), its coefficient grew larger, exceeding those of other variables (apart from the urbanicity), which mirrored observed patterns with chlamydia. This suggests that as residential segregation increases, populations living in highly segregated neighborhoods face a greater risk of gonorrhea infection. For the second period (2020–2021), the estimated coefficients for the residential segregation index followed the same patterns.

**Table 2.**
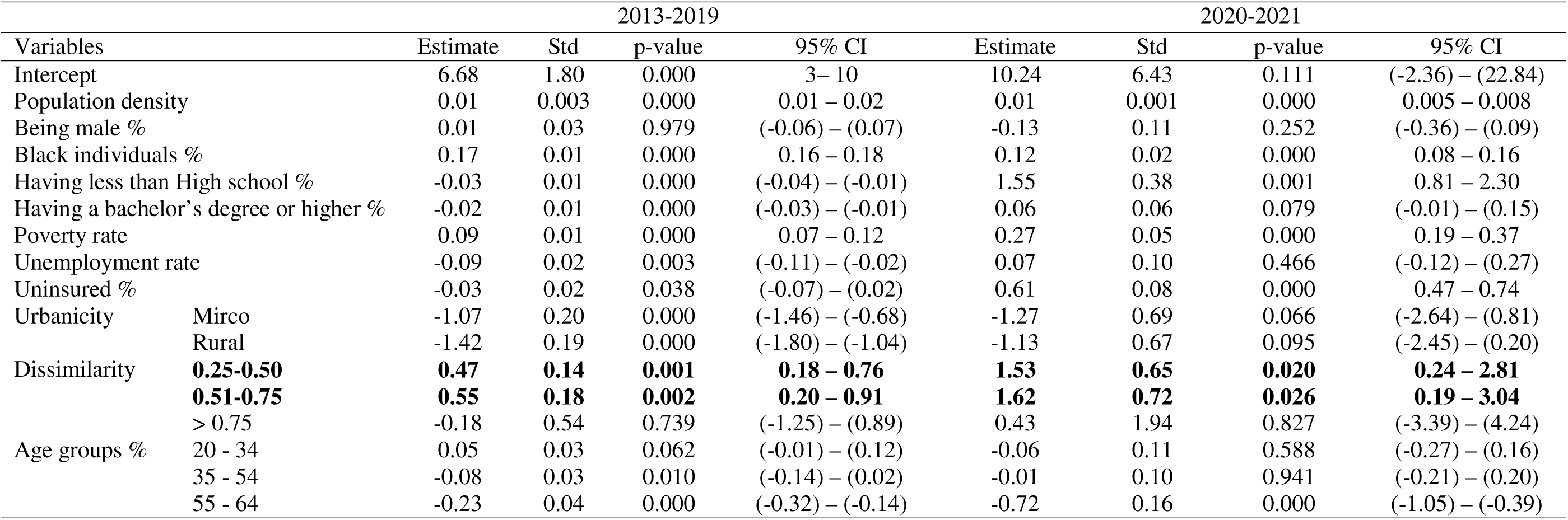
Generalized Estimating Equation’s result for gonorrhea from 2013 to 2021.

### 3.3. Spatial regression analysis

#### 3.3.1. Chlamydia

Figure 4 presents the spatial distribution of estimated coefficients for the association between racial segregation and chlamydia incidence rates across U.S. counties for the years 2013, 2014, and 2015. Due to space constraints, only these three years are displayed, while maps for other years are available in the supplementary material (Figure S3). The maps are divided into three groups based on segregation levels: Group 1 (0.25– 0.50, moderate segregation), Group 2 (0.50–0.75, high segregation), and Group 3 (>0.75, extreme segregation). Across all years, counties in states such as Mississippi, Alabama, Georgia, and South Carolina consistently exhibit strong positive associations in all three groups, particularly in areas with high and extreme segregation (Groups 2 and 3). Similarly, Texas, Louisiana, and Arkansas show moderate to strong associations, especially in highly segregated areas. In contrast, parts of the Midwest, including Minnesota, Wisconsin, and Iowa, display weaker or negative associations, particularly in Group 1 and Group 3. Additionally, some western states, such as Montana, Wyoming, and Utah, demonstrate spatial variability, with stronger positive associations in some counties and weaker associations in others. These patterns suggest that the relationship between racial segregation and chlamydia incidence is complicated and varies across different regions and segregation intensities.

**Figure 4.**
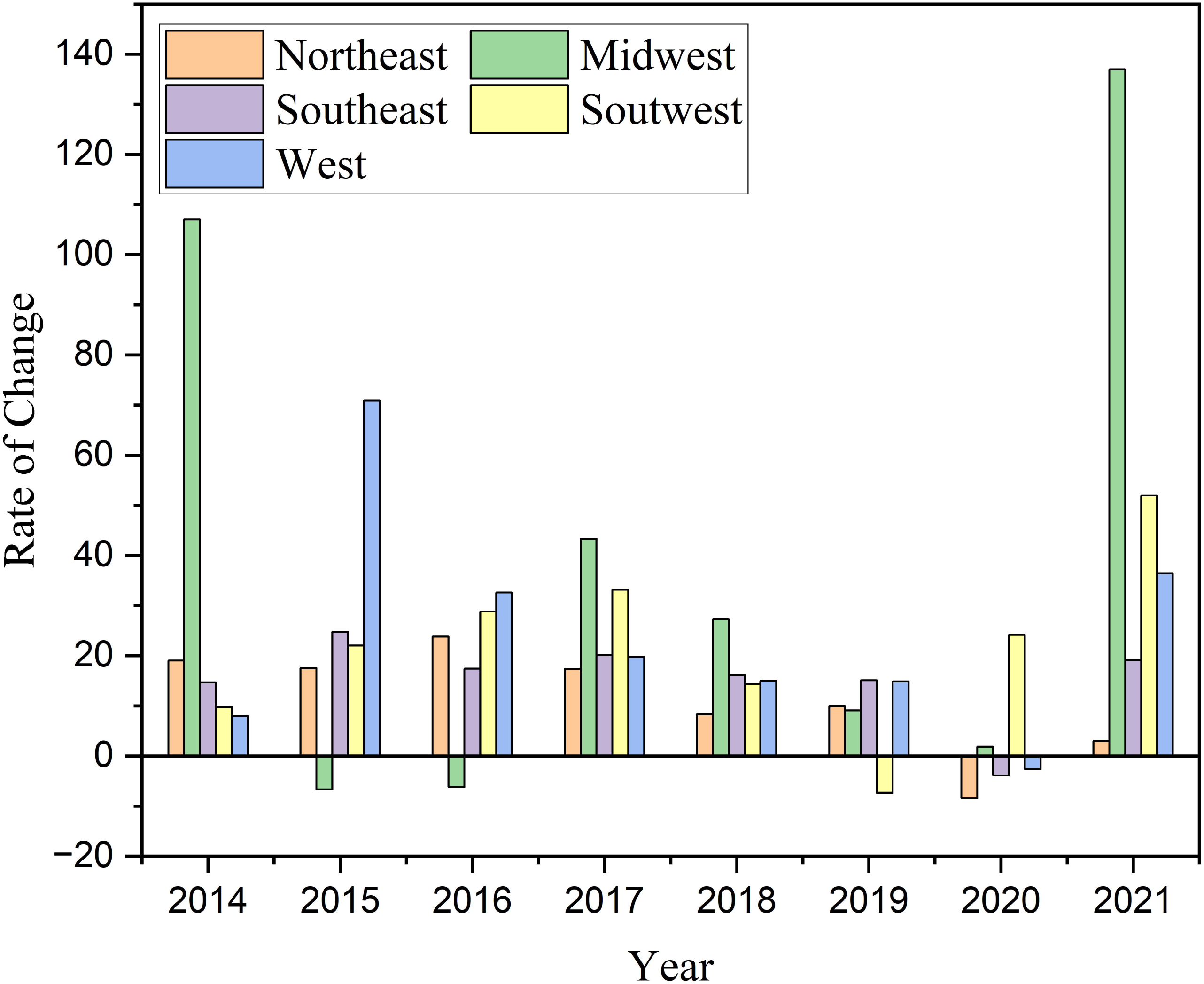
Segregation coefficient distribution for each year for chlamydia. The legend is displayed only on one map since the coefficient values were classified in the same range across all years to ensure a more comparable analysis.

#### 3.3.2. Gonorrhea

Figure 5 illustrates the spatial distribution of estimated coefficients for the relationship between racial segregation and gonorrhea incidence rates across U.S. counties for the years 2013, 2014, and 2015; the remaining years (2016-2021) is shown in the supplementary file (Figure S4). Similar to the chlamydia analysis, these maps are divided into three groups based on segregation levels: Group 1 (0.25–0.50), Group 2 (0.50–0.75), and Group 3 (>0.75). The spatial patterns reveal distinct regional variations. Across all years, counties in the Southeastern states, including Georgia, Alabama, Mississippi, and South Carolina, consistently exhibit strong positive associations across all three segregation groups. This suggests that in these regions, higher levels of residential segregation are significantly linked to increased gonorrhea incidence rates. Midwestern states, such as Illinois, Indiana, and Missouri, display varying patterns, with some areas showing strong positive associations, particularly in highly segregated counties (Groups 2 and 3). Western states, such as Texas, Arizona, and New Mexico, exhibit moderate associations, while Northern states, including Minnesota, North Dakota, and Montana, show weaker or even negative associations, especially in Group 1. Overall, the findings indicate that the relationship between residential segregation and gonorrhea incidence is more pronounced in the South and parts of the Midwest, while the associations are weaker in some Northern and Western regions.

**Figure 5.**
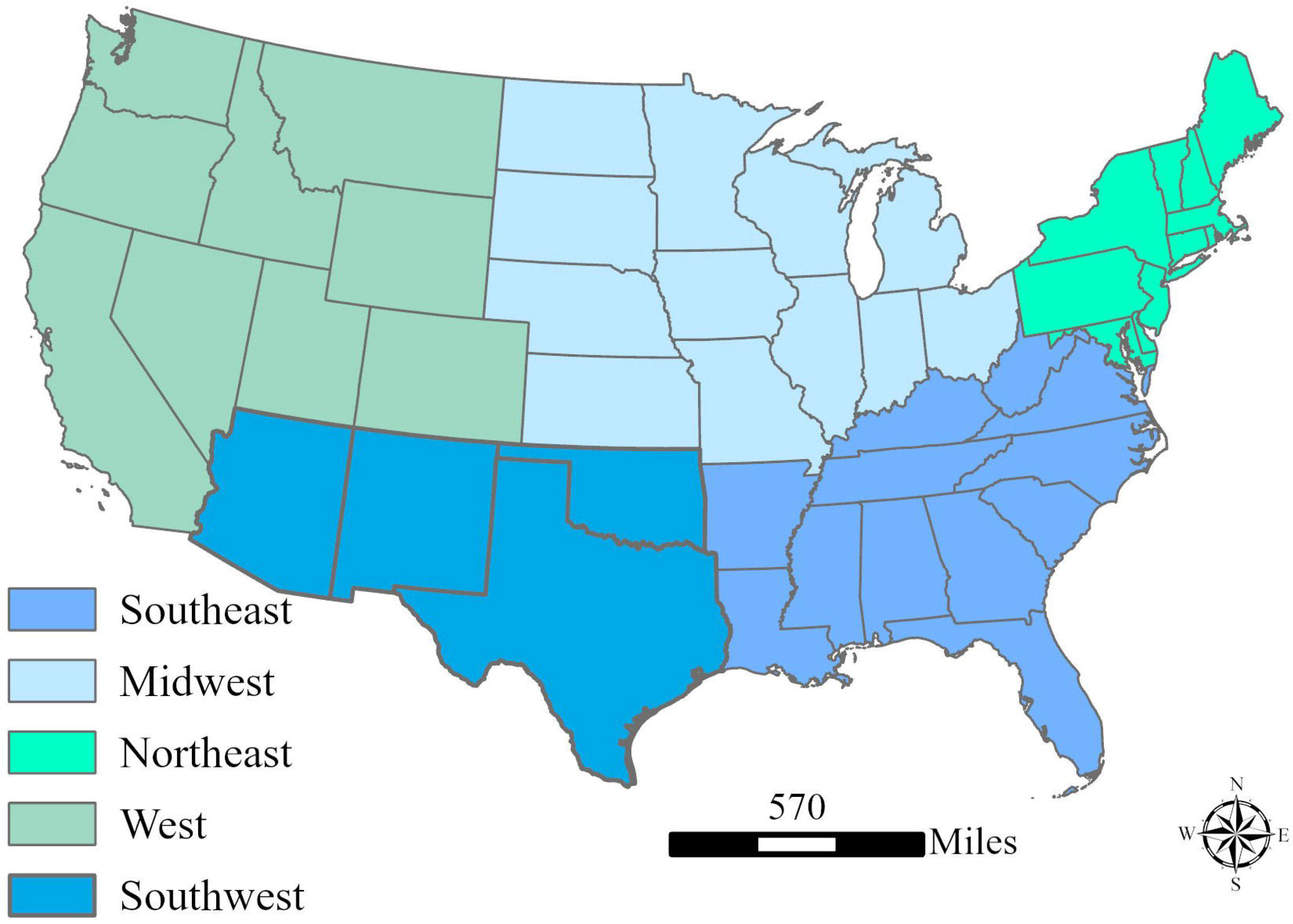
Segregation coefficient distribution for each year based for gonorrhea disease. The legend is displayed only on one map since the coefficient values were classified in the same range across all years to ensure a more comparable analysis.

## 4. DISCUSSION

This study analyzed the spatiotemporal patterns of chlamydia and gonorrhea incidence rates in the U.S. from 2013 to 2021, explored the relationship between the residential segregation and STI incidence rates (gonorrhea/chlamydia), and examined the impact of the COVID-19 pandemic on these associations. The geographic distribution of both infections varied across regions over time, predominantly affecting the Southeast and Midwest. Generally, the South, especially the Southeast, reported high incidence rates of chlamydia, while gonorrhea incidence rates were concentrated in the South and West. Chlamydia and gonorrhea trended upward between 2013 and 2021, but chlamydia experienced a decrease in 2020 and rebounded in 2021.

### 4.1. The influence of residential segregation on STI incidence rates

Our findings illustrate how residential segregation is significantly associated with chlamydia and gonorrhea transmission rates, underscoring the influence of structural inequities on disease acquirement. These results demonstrate that neighborhood segregation, particularly at moderate (0.25–0.50) and high (0.50-0.75) levels, was associated with higher incidence rates, suggesting that segregation may facilitate intergroup transmission among marginalized populations (21). However, extreme segregation (>0.75) generally showed weaker associations with the transmission of both diseases. These findings align with other findings within existing literature that highlight how systemic barriers such as reduced healthcare access, mass incarceration, and unemployment can lead to an increased risk of STI acquirement in predominantly Black neighborhoods (22). Indeed, the positive and large coefficient for Black individuals in both chlamydia and gonorrhea models support existing evidence that structural racism and its concentration of health depleting factors (e.g., poverty) exacerbates risks for STI acquirement (4). Furthermore, our results add to existing literature by suggesting that during COVID 19 pandemic, these disparities may have been further exacerbated, as many communities faced challenges like reductions in the number of services offered by STI service reduction, interruptions, and closures (23–25). As chlamydia is often asymptomatic, its diagnosis is heavily reliant on routine screening, which is significantly influenced by SDOH and was disrupted during the COVID-19 pandemic and potentially leading to underestimations of its incidence due to underdiagnosis. In contrast, gonorrhea typically presents symptoms, so its reported infection rate is more likely to reflect actual risk behaviors.

### 4.2. Geographic variety with the association between residential segregation and STI incidence rates

The findings from geospatial model underscored the persistent and spatially heterogeneous impact of residential segregation on the incidence rates of both diseases. In general, higher coefficients are commonly clustered in the Southeast and Midwest, with variations in the West and Upper Plains. This spatial stability over the study period indicates that residential segregation, particularly at moderate to extreme levels (≥0.25), is a consistent predictor of elevated gonorrhea and chlamydia transmission rates. Such patterns echo previous evidence linking structural inequalities like residential segregation and income inequality to adverse health outcomes (e.g., STI acquirement, infant mortality, pre-term birth) (26–28). It is also notable that the spatial distributions between residential segregation and chlamydia/ gonorrhea incidence varied over time. The complexity of spatial patterns and their changes highlight evolving socio structural drivers of gonorrhea incidence. Collectively, the spatial heterogeneity underscores the complex interplay between segregation and the transmission of chlamydia and gonorrhea. Therefore, addressing segregation-specific drivers of STI disparities requires the implementation of tailored and comprehensive interventions that take place at policy, institutional, and community levels (29–32).

### 4.3. The impact of COVID-19 on STI incidence and its association with residential segregation

Our results also suggest slight differences in how the COVID-19 pandemic impacted chlamydia and gonorrhea incidence rates. Chlamydia incidence rates declined noticeably during the pandemic. Gonorrhea incidence rates remained stable during the pandemic and experienced a sharp increase in the Midwest region of 136.95%. These findings align with the findings of Pollack et al. (2025). Increases in gonorrhea could be due to the resumption of regular STI testing services and demographic specific factors that fall outside the scope of this study (e.g., individual level behaviors) (26, 33). As such, additional research is needed to understand the mechanisms contributing to this increase in gonorrhea.

COVID-19 also had a significant impact on the associations between chlamydia incidence and sociodemographic factors. During the second period (2020–2021), the relationship between unemployment rate turned a significant predictor, highlighting the role of socioeconomic status in STI trends. Scholars like Woodhandler & Himmelstein (2020) conclude that employment-loss leading to health insurance changes can result in challenges like reduced healthcare access and delayed medical visits that exacerbate adverse health outcomes in disadvantaged populations (34–36).

Several risk factors of gonorrhea, including unemployment rate, micro-urban, and rural regions, became statistically insignificant during the pandemic (2020–2021). Existing literature notes that policies like lockdowns altered the sexual behavior of individuals, subsequently leading to fewer transmission opportunities, especially in lower-density areas. This could explain why we observed weakened associations between gonorrhea and micro-urban and rural settings during the pandemic. It is important to note that pandemic-related delays in STI surveillance and case reporting created backlogs, which could have resulted in potential underestimations of gonorrhea incidence and thus potentially impacting statistical relationships. Therefore, our pandemic-specific results should be interpreted with caution. Despite the disruptions caused by COVID-19 pandemic, the association between residential segregation and STI incidence (i.e., chlamydia and gonorrhea) remained positive and statistically significant, particularly for moderate and high segregation levels.

### 4.5. Limitations

Despite this comprehensive analysis using temporal and spatial regression models, several limitations remain. First, the dataset consisted of 3,058 counties within the contiguous United States. As a result, our analysis does not take into account how segregation may influence chlamydia and gonorrhea incidence rates within non-contiguous states. Second, COVID-19 disrupted the study period, producing a notable decline in chlamydia incidence rates and a minor decrease in gonorrhea incidence rates, possibly skewing results. Future investigations should extend beyond this timeframe to investigate if STIs truly decreased or continued trending upward post-2020. Finally, we only considered Black individuals within our segregation variable, excluding other racial/ethnic groups. Incorporating additional demographics could offer deeper and more nuanced insights.

## 5. CONCLUSION

This study examined the relationship between chlamydia, gonorrhea, and residential segregation from 2013-2021 in the United States. Our findings add to existing literature through revealing how residential segregation remained a substantial driver of chlamydia and gonorrhea transmission during this period, underscoring the influence of structural inequities on STI incidence rates. It is important to note that, due to the sexual healthcare service interruptions from the COVID-19 pandemic in tandem with the decision to only include 2021 after the COVID-19 pandemic within our analysis, these findings should be interpreted with caution. Future studies should examine the relationship between STIs and racial segregation with populations outside of White and Black populations. Additionally, future studies should assess how residential segregation influences the incidence rates of other sexually transmitted infections (e.g., syphilis).

## ABBREVIATIONS

(STI): Sexually Transmitted Infections
(SDOH): Social Determinants of Health
(CDC): Centers for Disease Control and Prevention’s
(USDA): U.S Department of Agriculture
(SGWR): The Similarity and Geographically Weighted Regression

## Data Availability

All data used within this study were obtained from publicly available secondary data sources. Therefore, no new data were generated by the authors for the present study.

---

**Figure S1:**
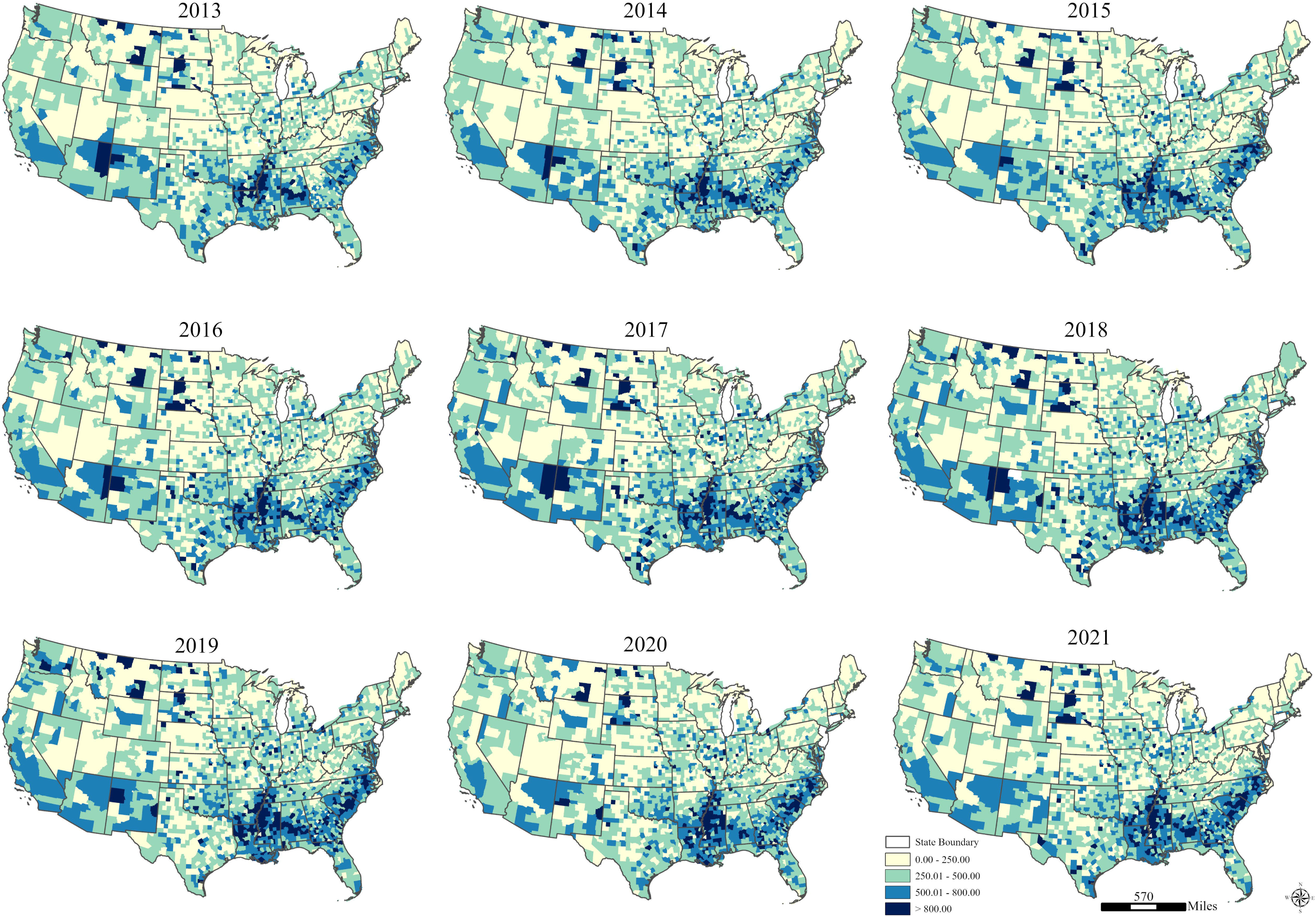
The temporal distribution of chlamydia disease at the state level.

**Figure S2:**
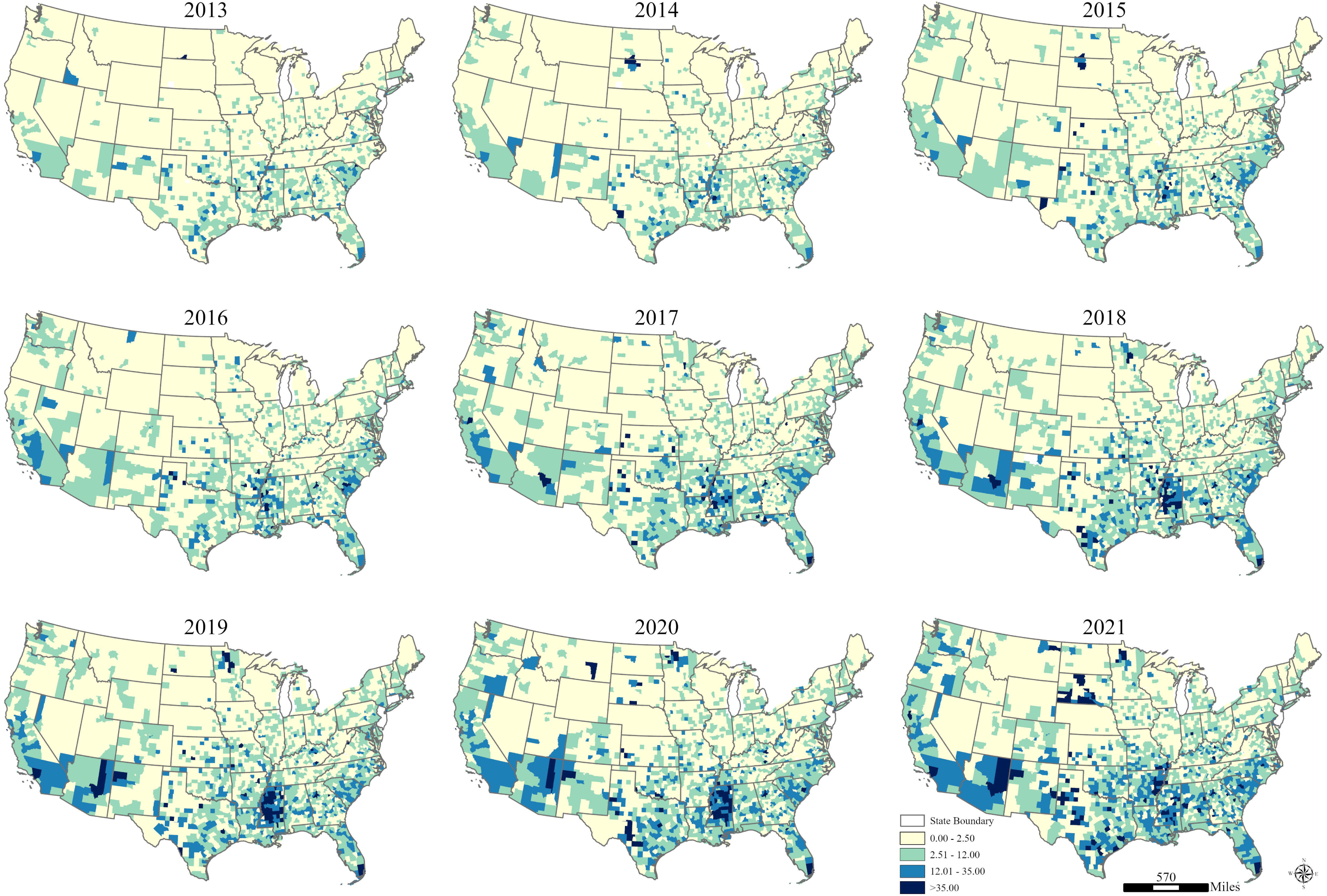
The temporal distribution of gonorrhea disease at the state level.

**Figure S3:**
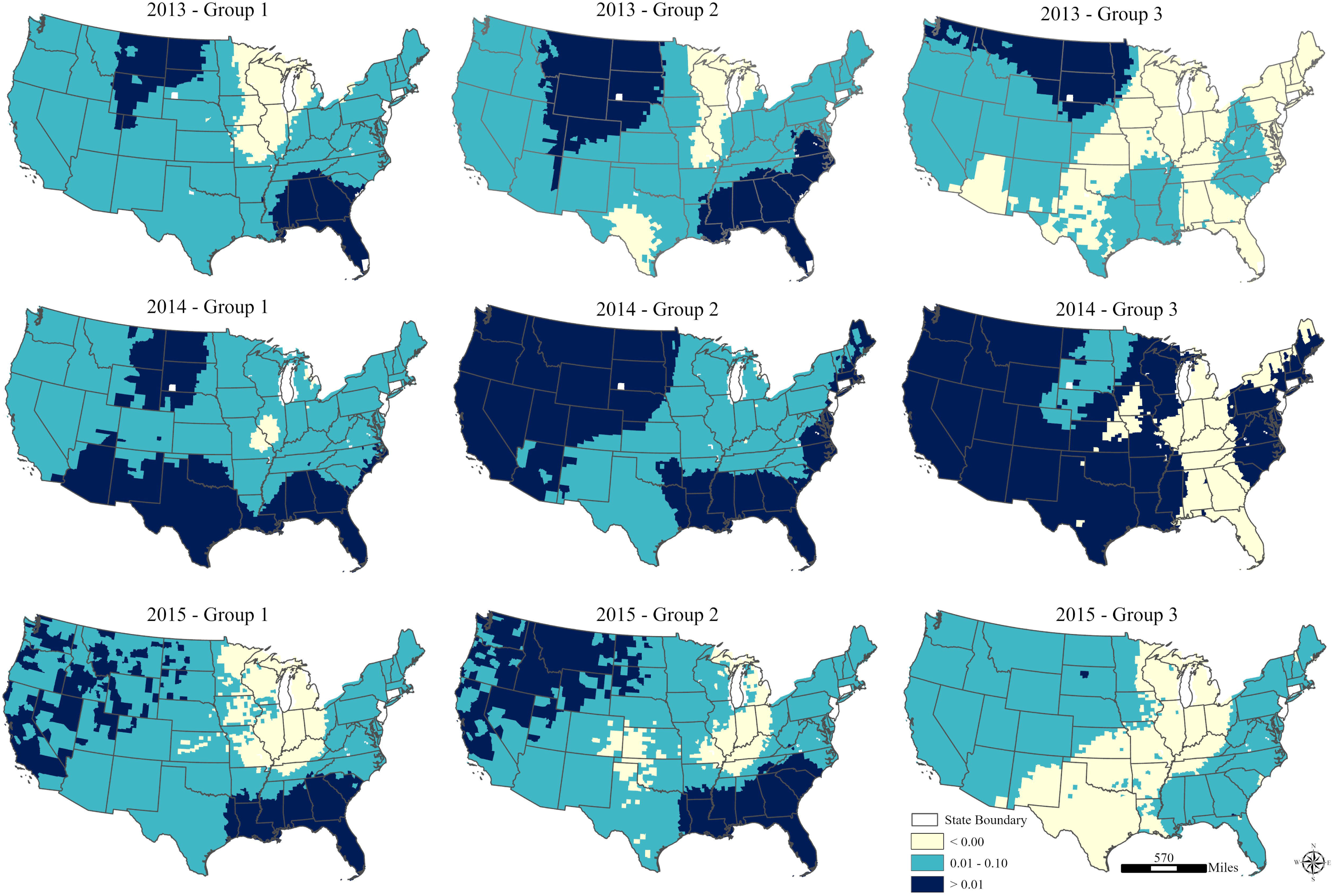
Coefficient of dissimilarity index (segregation index) from 2016 to 2021 for chlamydia illness.

**Figure S4:**
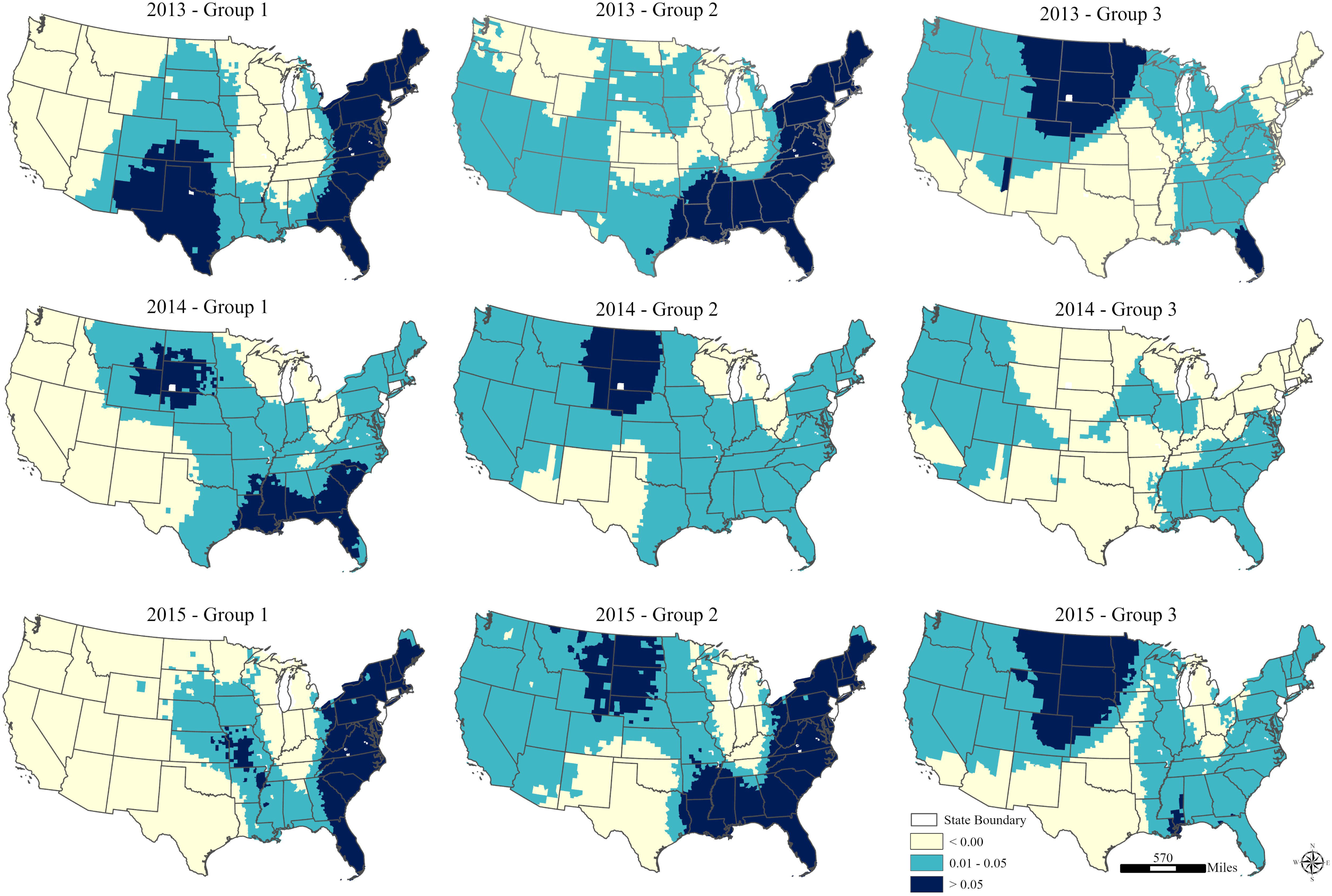
Coefficient of dissimilarity index (segregation index) from 2016 to 2021 for gonorrhea illness.

