## Supplementary figures and images for "Chlamydia and Gonorrhea Incidence and Residential Segregation: US Spatiotemporal Patterns (2013–2021)"

### Figure S1.jpeg

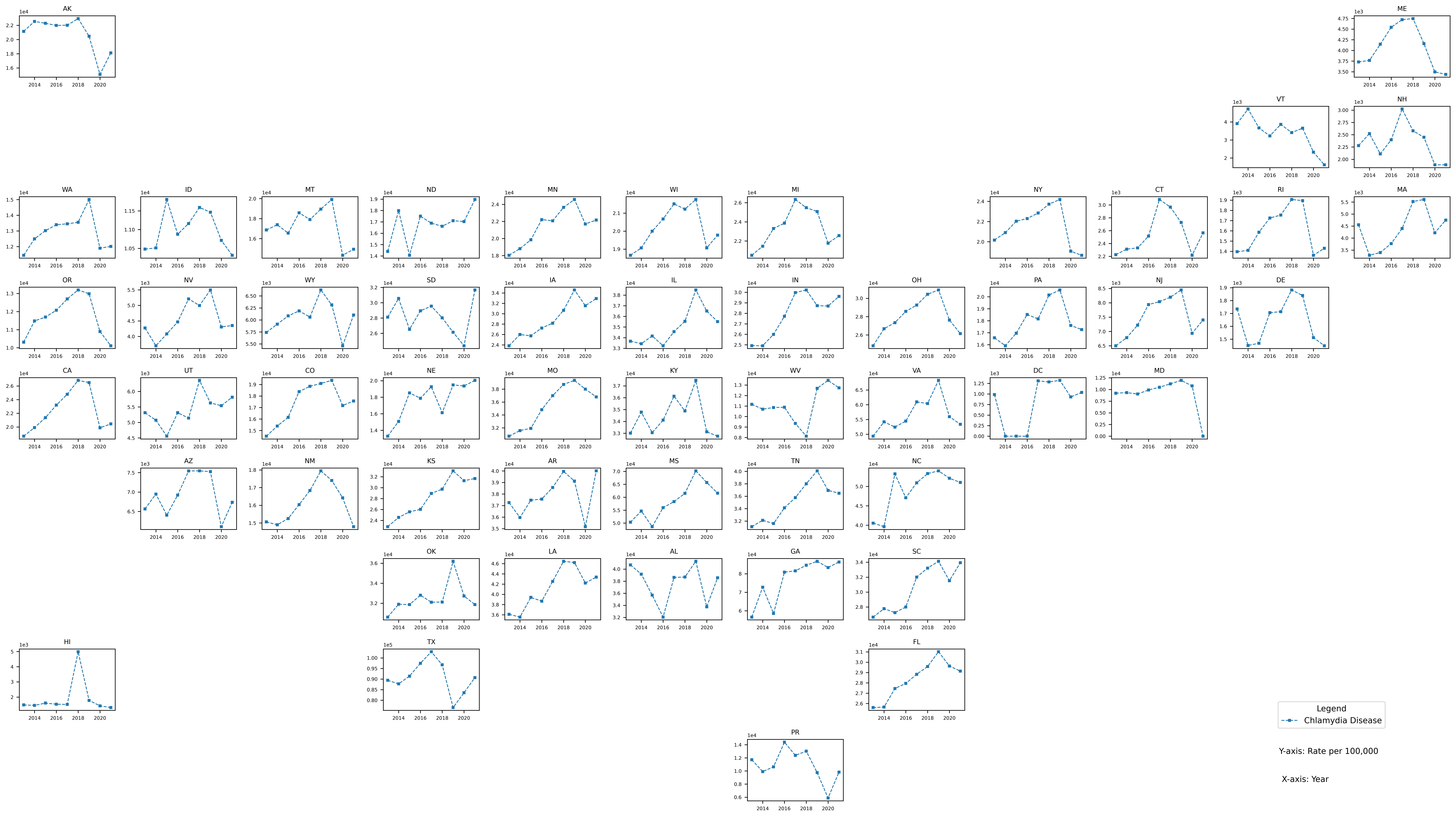

### Figure S2.jpeg

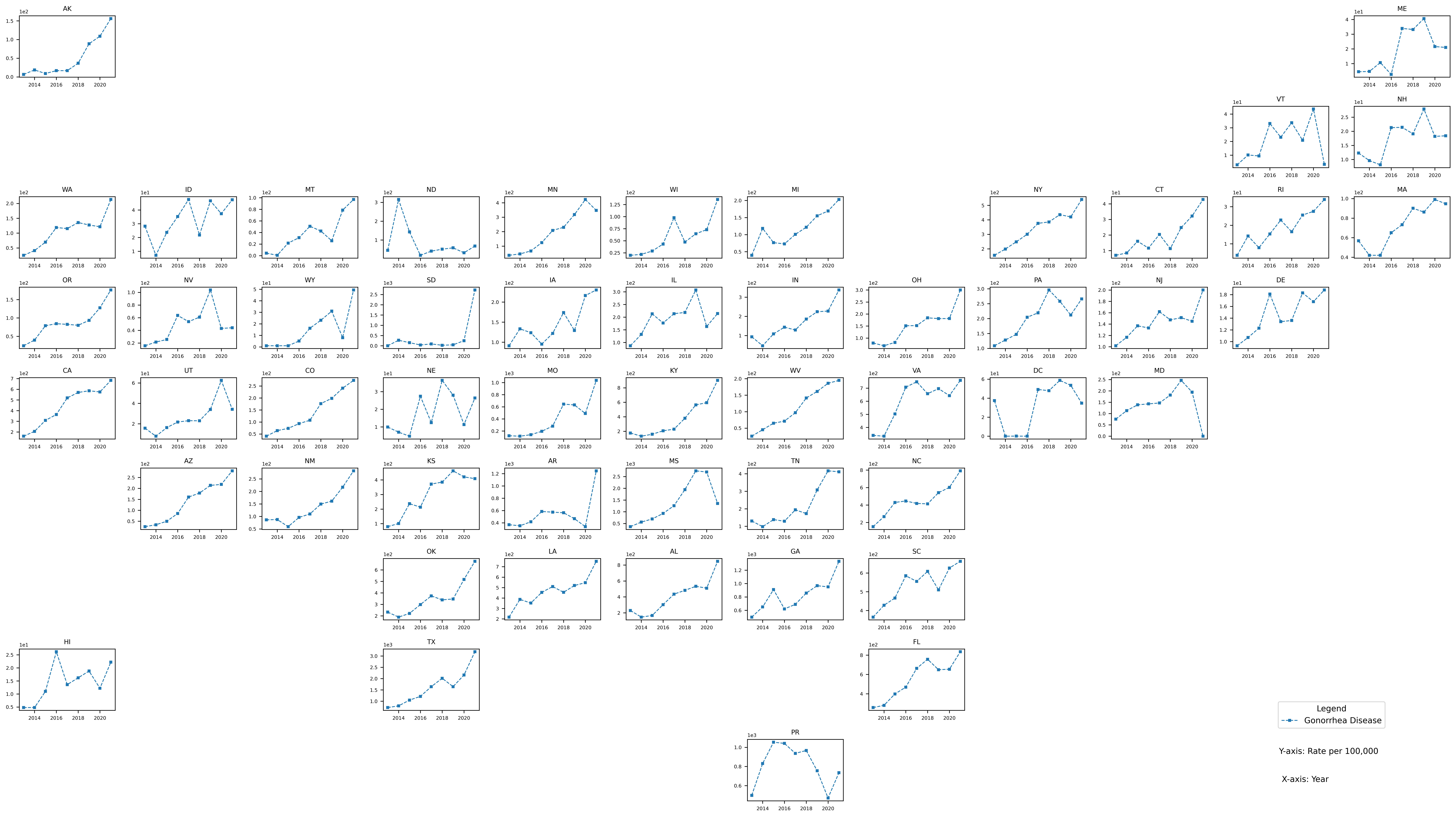

### Figure S3.jpeg

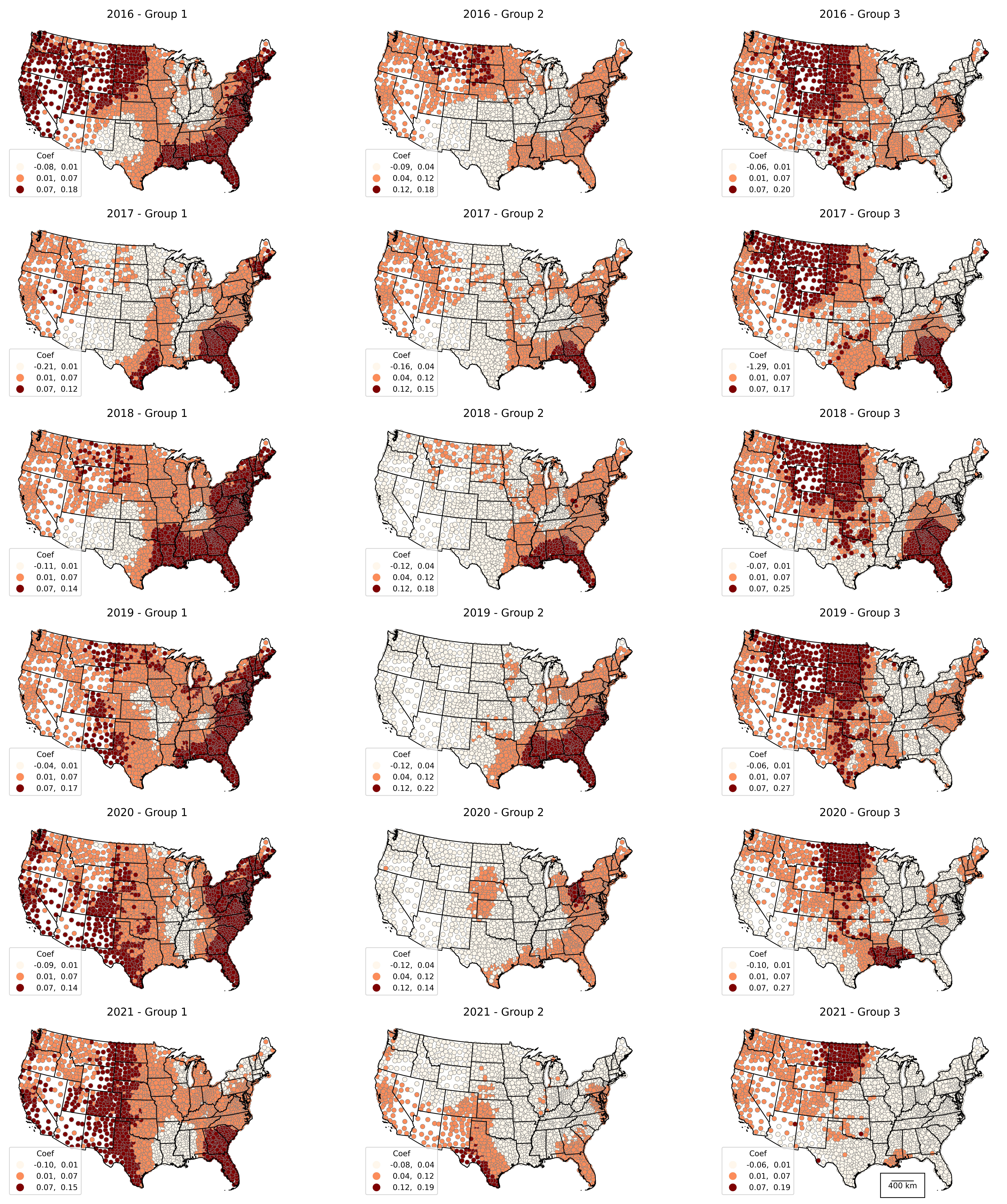

### Figure S4.jpeg

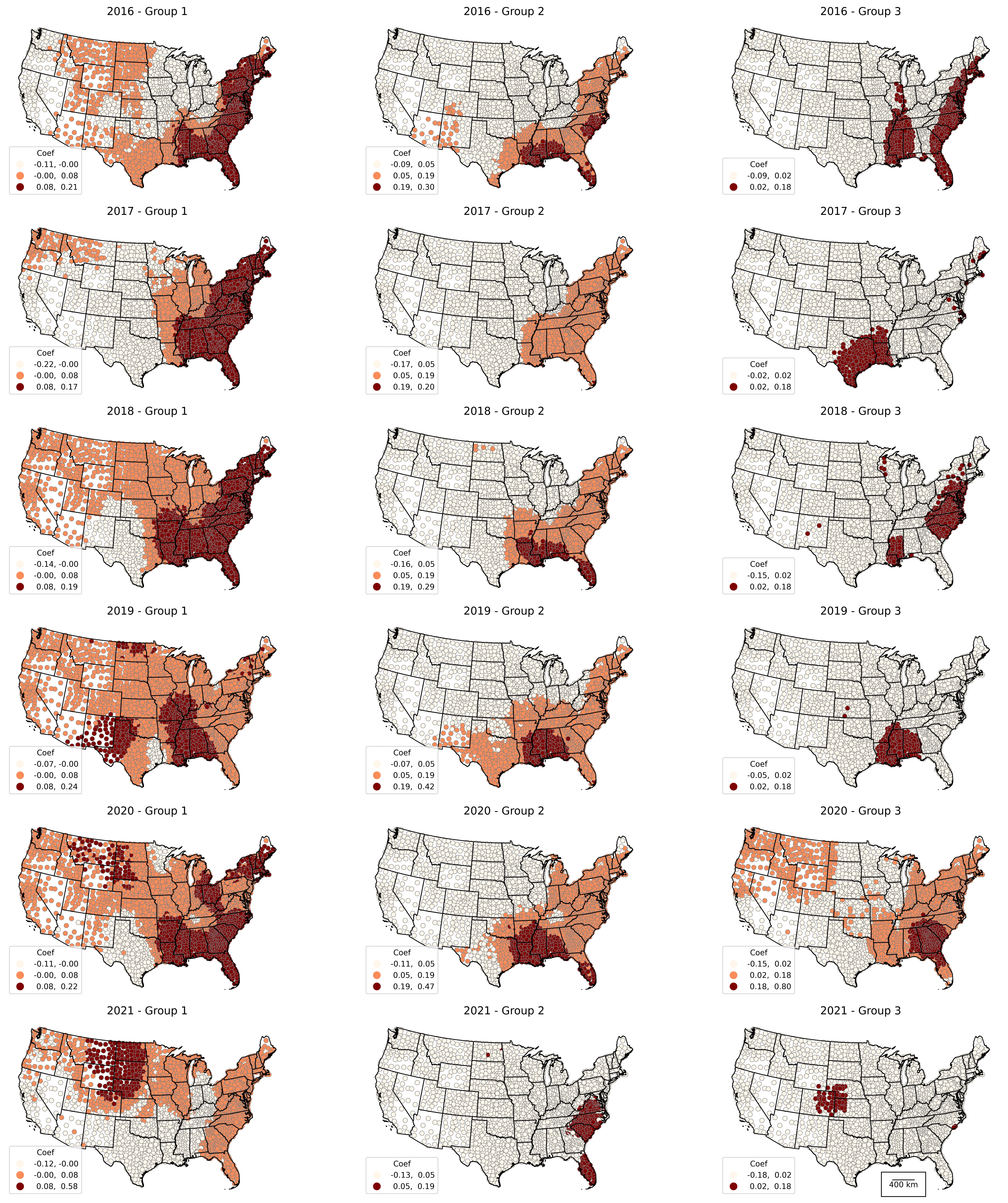
